# Hybrid-Quantum approach for the optimal lockdown to stop the SARS-CoV-2 community spread subject to maximizing nation economy globally

**DOI:** 10.1101/2021.06.14.21258907

**Authors:** Sahil Zaman, Alex Khan, Arindam Sadhu, Kunal Das, Faisal Shah Khan

## Abstract

Owing to the SARS-CoV-2 epidemic (severe acute respiratory coronavirus 2 syndromes), the global situation has changed drastically. Several countries, including India, Europe, U.S.A., introduced a full state/nation lockdown to minimize the disease transmission through human interaction after the virus entered the population and to minimize the loss of human life. Millions of people have gone unemployed due to lockdown implementation, resulting in business and industry closure and leading to a national economic slowdown. Therefore, preventing the spread of the COVID-19 virus in the world while also preserving the global economy is an essential problem requiring an effective and immediate solution. Using the compartmental epidemiology S, E, I, R or D (Susceptible, Exposed, Infectious, Recovery or Death) model extended to multiple population regions we predict the evolution of the SARS-CoV-2 disease and construct an optimally scheduled lockdown calendar to execute lockdown over phases, using the well-known Knapsack problem. A comparative analysis of both classical and quantum models shows that our model decreases SARS-CoV-2 active cases while retaining the average global economic factor, GDP, in contrast to the scenario with no lockdown.

## 1. Introduction

The cause of ongoing health threat to almost every country of the world is the severe acute respiratory syndrome coronavirus 2 (SARS-CoV-2). Mostly it is known as COVID-19 (CoronaVirus Disease 19). Almost 32.4 million people worldwide have been affected by the COVID-19 virus as of September 2020. Whereas, 0.98millionhave died of COVID-19 globally, and 0.22million people have recovered from this disease. We know that this deadly virus spreads from one person to another through tiny droplets of the virus from COVID-19 affected persons who exhale or cough. Again, these microdroplets can stick to metals or any exterior object for a limited time. Others can become infected by touching these COVID-19 infected objects or surfaces if they further touch their nose and mouth. Other shared devices and objects have also been known to transmit the disease. Same way, anybody can also catch COVID-19 if they breathe in droplets from a COVID-19 infected person, coughs out, or exhales droplets [1-2]. All these contribute to the community spread of the disease. This is why it is vital to maintain social distancing, home quarantine, lockdown, frequent hand washing, etc. to minimize the effect of the COVID-19 virus.

As the vaccine for the deadly virus COVID-19 still is not available, many researchers are involved in finding a suitable mathematical model that will forecast this epidemic. Many scientific epidemiological studies are reported from 1927 to present pandemic situations [3-8]. In the article [3], Kermark et. al. presented the compartmental modeling in 1927. From then on, many epidemic mathematical models are reported in [4-8] like SIR, SIRD, SEIR, *θ*-SEIHRD, etc. Ivorra, Benjamin et. al. presented their mathematical model in [8] and they include undetected infection and requirement of hospital beds. Nevertheless, all these models are specified with a fixed population of an area. These models do not address the evolution of disease in different population areas through travel or transfer of the disease between the areas.

The effects of travel related to many epidemiological mathematical models is introducedin [9-20]. In articles [9,10], Ahmed et al. and Sattenspiel et. al. address travel among the population, where the travel can be easily quantified. Another model regarding the spread of disease over multiple patches are discussed in Ding et al. [11] and Sattenspiel et al. [12, 13]. The SEIR and SEIRS model are discussed with their constant immigration in multipatches [15-16]. Again, Wang et. al. also added the quarantine concept to restrict the travel of infected persons in [16]. Nevertheless, all models ignore the transmission of diseases during travel. An epidemic SIS model with steady-state analysis is proposed by L. J. S. Allen et. al. in [17] where the transport-related infection is included. Again, all models identify the common solution to minimize the effect of COVID-19 as a lockdown. But constant lockdown creates a huge impact on GDP growth, which is not addressed in the case of a long term and on-going situation as we are seeing now.

Hence, we have to concentrate on the formation of a mathematical model that maintains a balance between the spread of COVID-19 and GDP growth. Our model is only a starting point and we used a limited graphical representation and duration to show that it is viable.

The manuscript is arranged as follows. In Section 2 the proposed multi-city SEIRD model with a transfer function is defined. In section 3 we are discussed about computing configuration. The result and discussion part are illustrated in section 4. And finally, the conclusion is found in section 5.

## 2. Methods and Materials

### 2.1) Mathematical Formulation

To determine the optimal lockdown schedule, we estimate the virus course over time using the five compartments of the mathematical epidemiology classical model SEIRD (Susceptible, Exposed, Infected, Recovered, and Death):

**S:** Number of susceptible individuals likely to get infected with the virus. The susceptible individual discovers the disease upon contact with the infected person and transfers to the exposed compartment.

**E:** Number of individuals that were exposed. There is a substantial time of incubation during which the persons become infected but are not yet infectious. At this time the person is in an Exposed compartment.

**I:** Number of infected individuals the exposed persons remain contagious after the incubation phase and can infect susceptible people.

**R:** Number of infected individuals who survived (and immune) from the disease and entered the recovered compartment. Cannot reinfect these susceptible individuals.

**D:** Number of deaths that have occurred as a result of the disease.

The SEIRD model works on one area or city, to make it a multicity-SEIRD model, we added a transfer function with the mathematical formulation as shown in figure 1. The novel transmission model spreads the virus over time into various cities. Hence the proposed formulations of susceptible rate, exposed rate, and infected rate are expressed in equations 1, 2, and 3 respectively. Whereas, equation 4 and 5 illustrates the rate of recovery and death respectively.

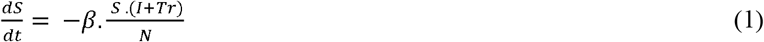

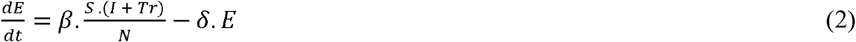

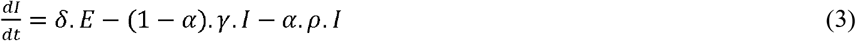

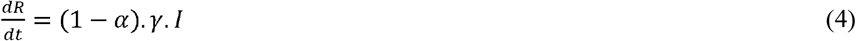

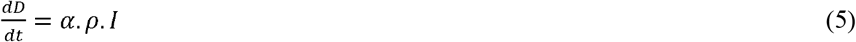

**Fig 1:**
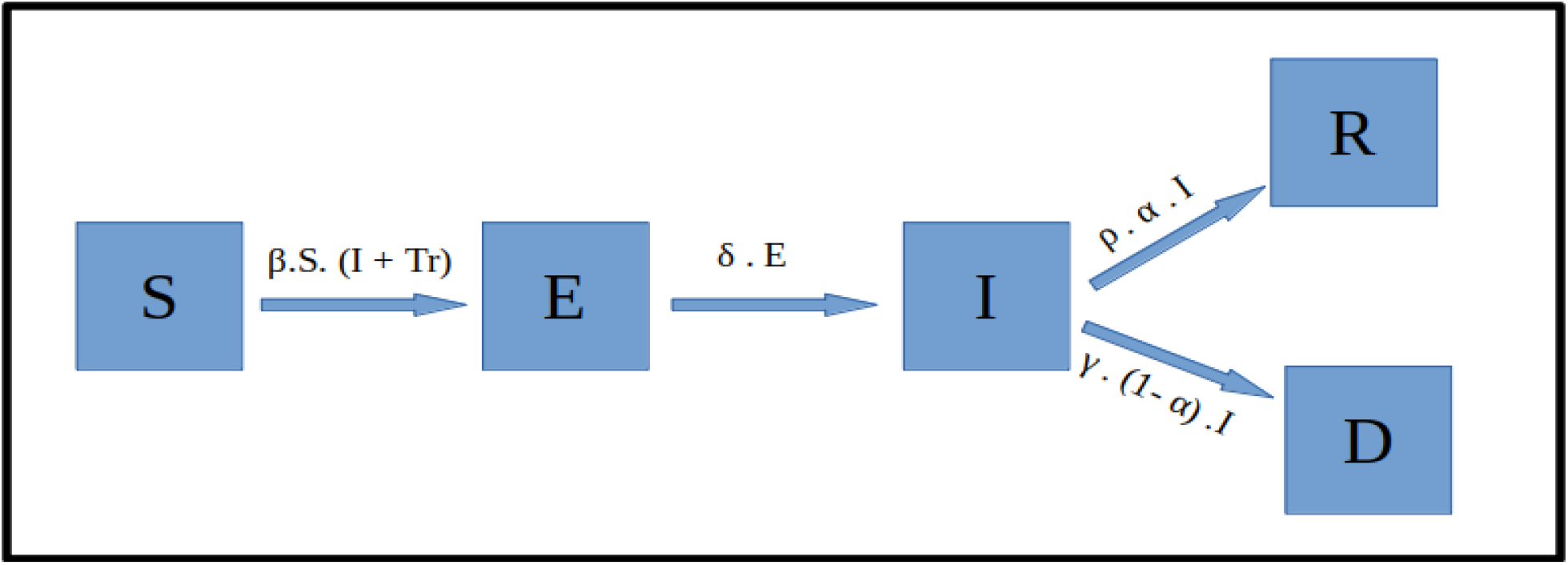
Multicity-SEIRD Model. Blocks represent the compartments, arrows represent individual flows between compartments over time, and parameters above the arrows valued.

### 2.2) Transfer Function

The virus spreads by communication between infected and susceptible persons. Hence, the intracity contact ratio is defined by *β* while the intercity is defined by *Tr* which depends on the distance between two cities and the individual traveling from one city to another. Let, *I*_*A,B*_ and *I*_*B,A*_ the number of infected populations traveling from city A to B and vise versa respectively as depicted in figure 2. Assuming that the total population of a city remains constant over time i.e. *I*_*A,B*_ ≈ *I*_*B,A*_, which in turn does not affect the property of the SEIRD model, i.e *N* =*S*(*t*) + *E*(*t*) + *I*(*t*) + *R*(*t)* + *D*(*t*), which states that the total population of a city remains constant, but only transmits the virus into the city affecting the *β*. The distance matrix represents the distance between two cities, where the linear term is 0 and the quadratic term, *d*_*A,B*_, *A* ≠ *B* is the distance. If the distance increases, the spread of the disease decreases, but the spread depends on several factors. Hence, we introduced a calibration constant *k*. Thus, the Tr function is defined as in equation 6

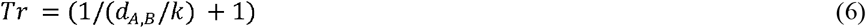

**Fig 2.**
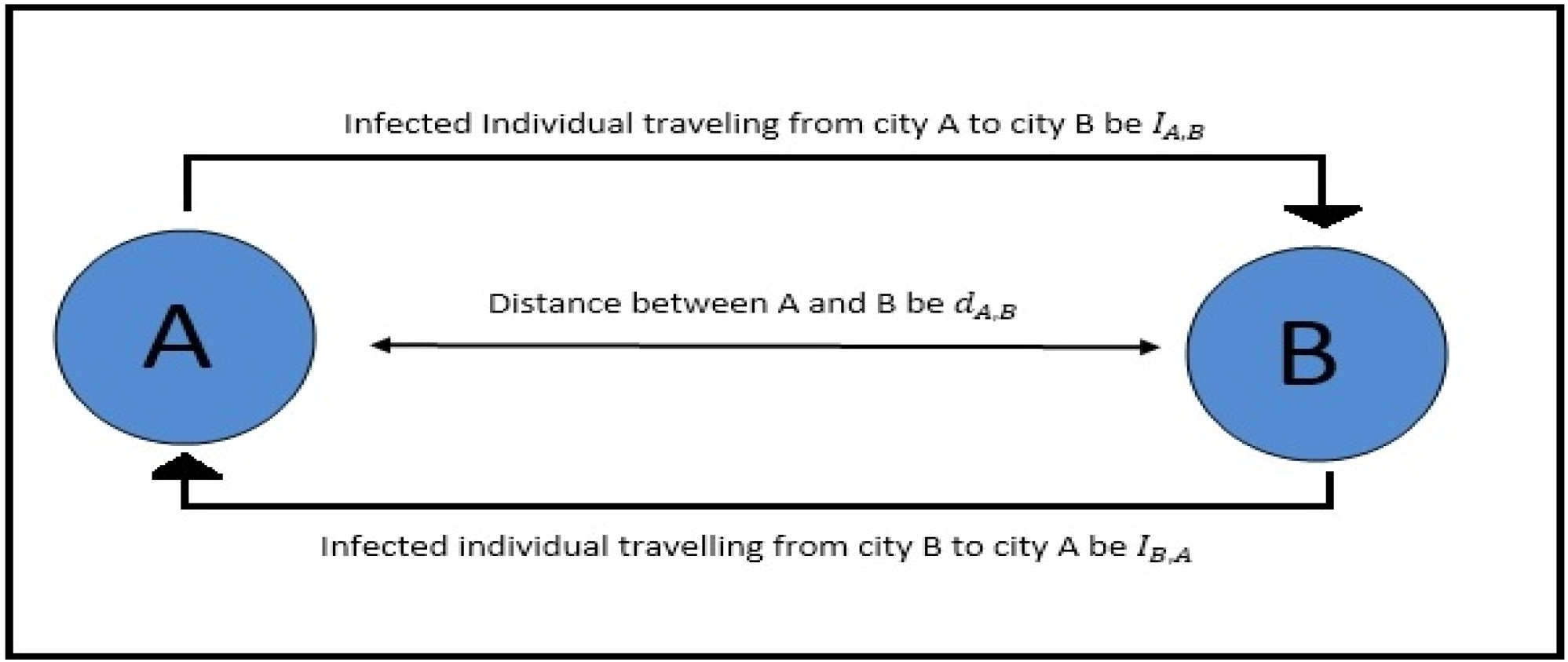
Transfer Function. The transmission of the virus is shown between two cities, A and B, where is the number of individuals traveling between the cities. The distance between A and B is.

We assume that the effect of one city’s infected population on other falls as the inverse of the distance. Other exponential decay functions can also be used and calibrated here. The key point is to model the effect of one city’s infected population on the other based on the movement of people between the cities. This can be through various means of transportation that connect the cities. The overall function can be calibrated based on real data by adjusting the calibration constant k. As *k* is reduced the impact decreases rapidly with distance as depicted in figure 3.

**Fig 3:**
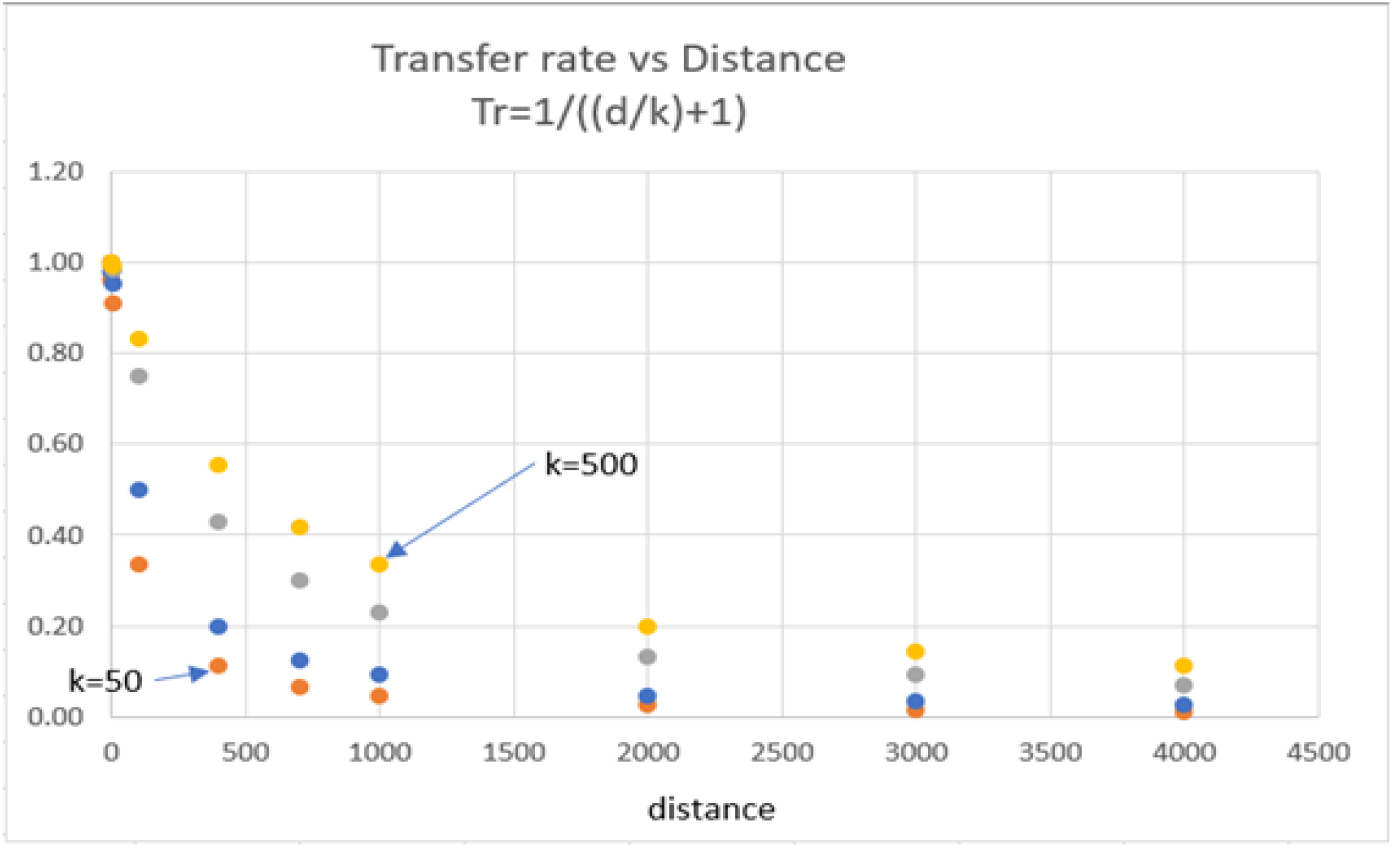
Transfer rate vs Distance graph for different values of k.

**Fig 4:**
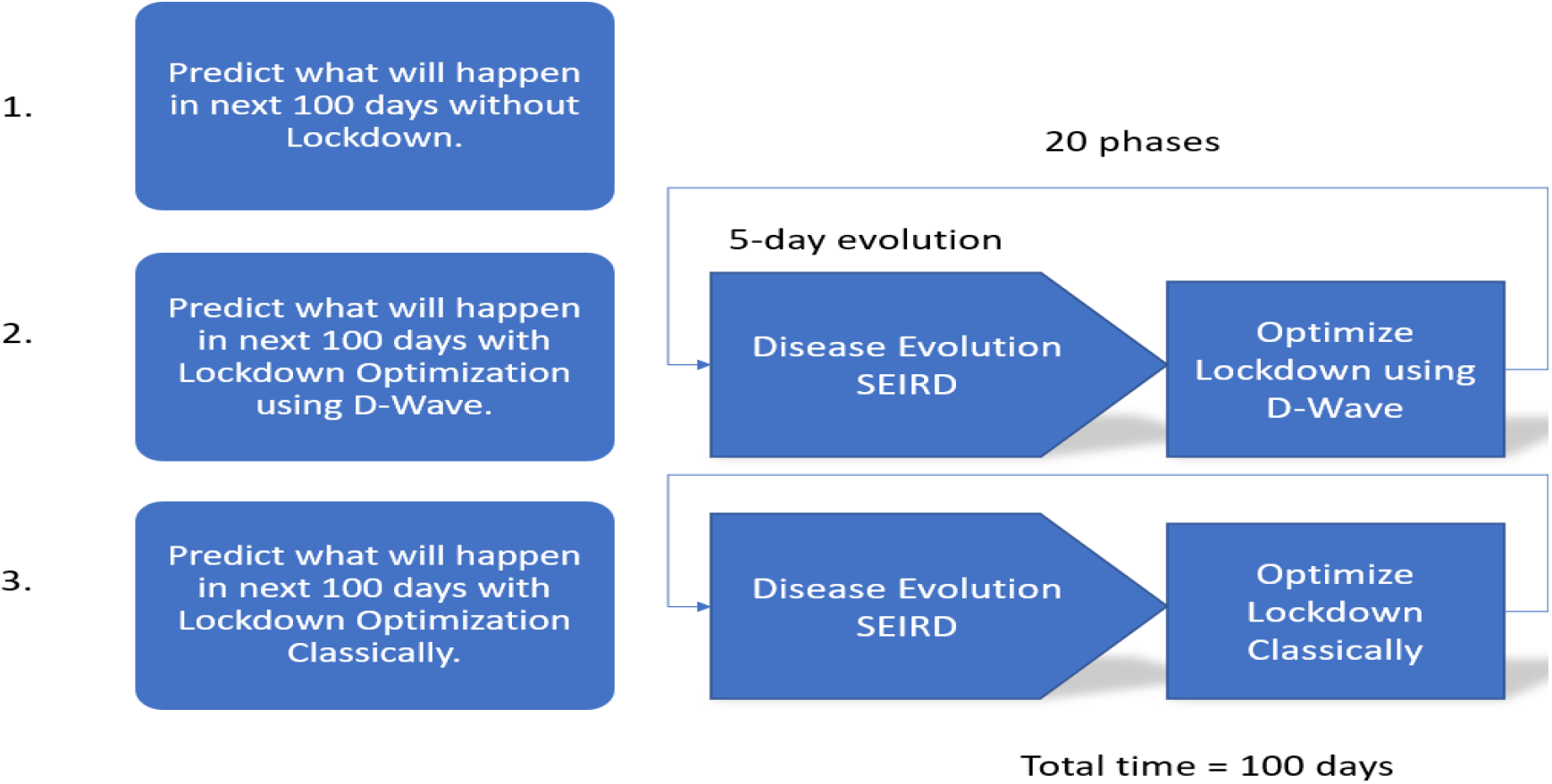
Flow of work. Initially without lockdown and then using the knapsack formulation results are derived from the quantum computer and classically.

### 2.3) Lockdown Optimization

To optimize a lockdown schedule, it is important to create an objective function which balances the conflicting dynamics of the need to close a city to reduce the spread vs the need to open a city to maintain an economy and thus GDP. We consider the objective function at a high level to be the statement below

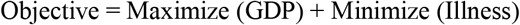

This objective is converted into an energy function in the form of a QUBO which can then be minimized both classically and using D-Wave Quantum Annealer.

In this project, we use Knapsack Quantum Algorithm to find the Optimized Lockdown subject to maximizing the GDP globally. The quantum algorithm for the knapsack problem is formulated by Andrew Lucas et. al. as reported in [21]. As per the knapsack problem, we want to minimize the weight that means the number of infected populations and the subject to maximize the profit that is GDP in our case. The knapsack formulation is expressed in equation 7.

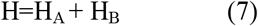

Where

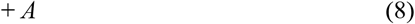

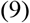

Where H_A_in equation 8, states the inequality of minimizing the weight of the knapsack, and H_B_in equation 9, states to maximize the total profit of knapsack. For our case objects that the open cities are in a knapsack, and the object or cities that are not included in knapsack means that the cities are in lockdown for the specified phase. Hence Open cities have a running economy means that open cities can utilize their hospital capacity (knapsack) to maintain. Whereas the economy for the closed one is assumed to be zero. Hence using the knapsack QUBO, we can find out a solution to our problem that is optimized lockdown subject to maximizing the GDP globally.

## 3. Computing Configuration

Using our formulations, we ran the model for 100 days without lockdowns for several cities. Which gives us the worst-case scenario of all the city over time due to the virus. An open city has a higher contact ratio and R0 than the state which is a lockdown. We split the total time into smaller phases, assuming for a phase, the state will stay locked down/ open. Over each phase, using the Multicity-SEIRD Model, we implement the optimal lockdown over cities using classical and Quantum Knapsack Problem, which will affect the movement of the virus through multiple cities for the next phase.

### 3.1) Model 0: No Lockdown

We took four states namely “Mumbai”, “Delhi”, “Chennai”, “Kolkata” and “Bengaluru”. The initial status of our experimental model is given in Table 1, where only the state “Mumbai” has an Exposed population of 100. The bed capacity is the estimated bed available for the infected population of COVID-19 and the GDP of each state in the U.S. Dollar. The total bed size is 802213 and the overall GDP is 906.8. The constant parameter for the Multicity-SEIRD model, given in Table 7 (APPENDIX), are experimental arbitrary values. Contact Ratio: β = 1.25, Incubation period: δ = 0.2, Death rate: α= 0.02, proportion of infected recovering per day: γ = 0.25, total number of people an infected person infects: R0 = 5.0, number of days an infected person has and can spread the disease D^*^ = 4.0, days from infection until death: ρ = 7.0.

**Table 1.** Initial condition of five states

| Index | State | Total Population | Exposed | Bed capacity | GDP (USD) |
| --- | --- | --- | --- | --- | --- |
| 1 | Mumbai | 122153000 | 100 | 231739 | 310.0 |
| 2 | Delhi | 19814000 | 0 | 155375 | 293.0 |
| 3 | Chennai | 75695000 | 0 | 262109 | 78.6 |
| 4 | Kolkata | 96906000 | 0 | 39455 | 150.0 |
| 5 | Bengaluru | 11883000 | 0 | 113535 | 45.2 |

The Transfer function Tr use the distance matrix, (element is the distance (in Km) between states i and j), and calibrating constant k = 2.

For the initial run of the Multicity-SEIRD Model, without implementing any lockdown to any of the five states for 101 days, we get the result as follows.

The plotin Fig 5 show the normal evolution of a disease through a population. The plot without lockdown shows that disease is spread quickly from the initial population center where we started with 100 infected individuals to other areas. The disease then grows exponentially through the population unconstrained and causes a considerable rise in the infected population which would need to be hospitalized. However, without any lockdown measures, this quantity would overwhelm the healthcare infrastructure and cause considerable devastation. As the disease evolves further the susceptible population eventually goes down to zero, while the recovered and dead populations reach their maximum. This would be the opposite of a full lockdown or “herd” immunity where those that are naturally able to ward off the disease would recover and there would be considerable deaths.

**Fig 5:**
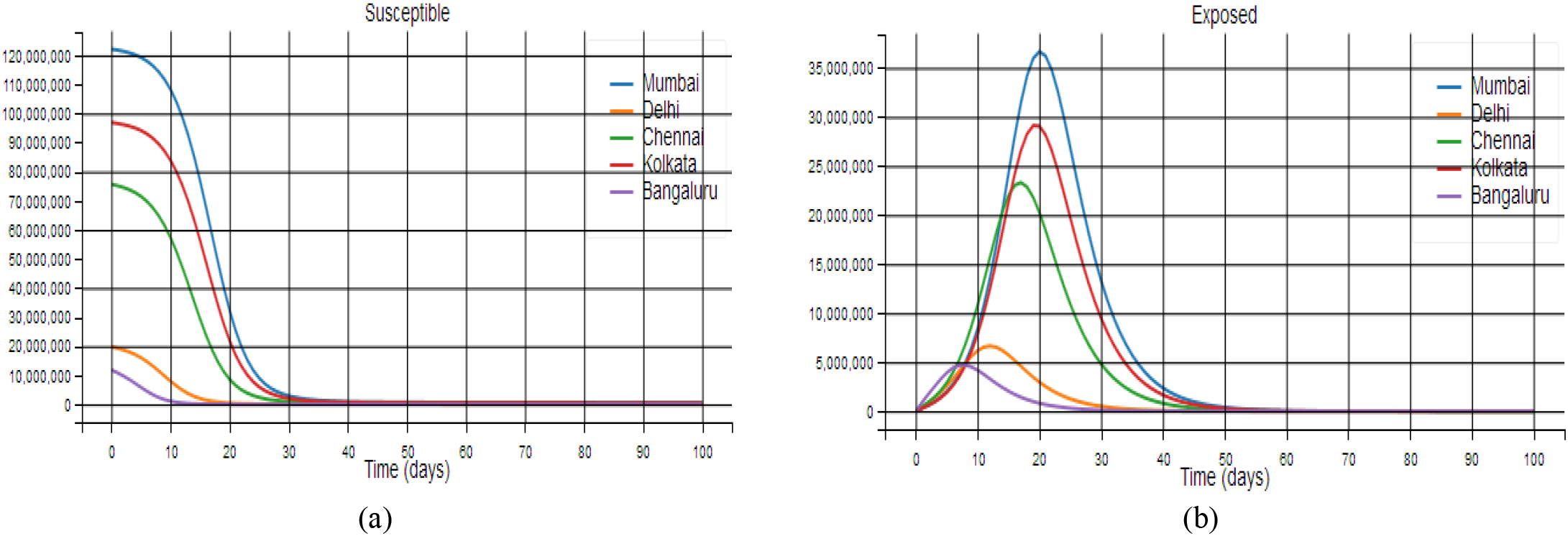
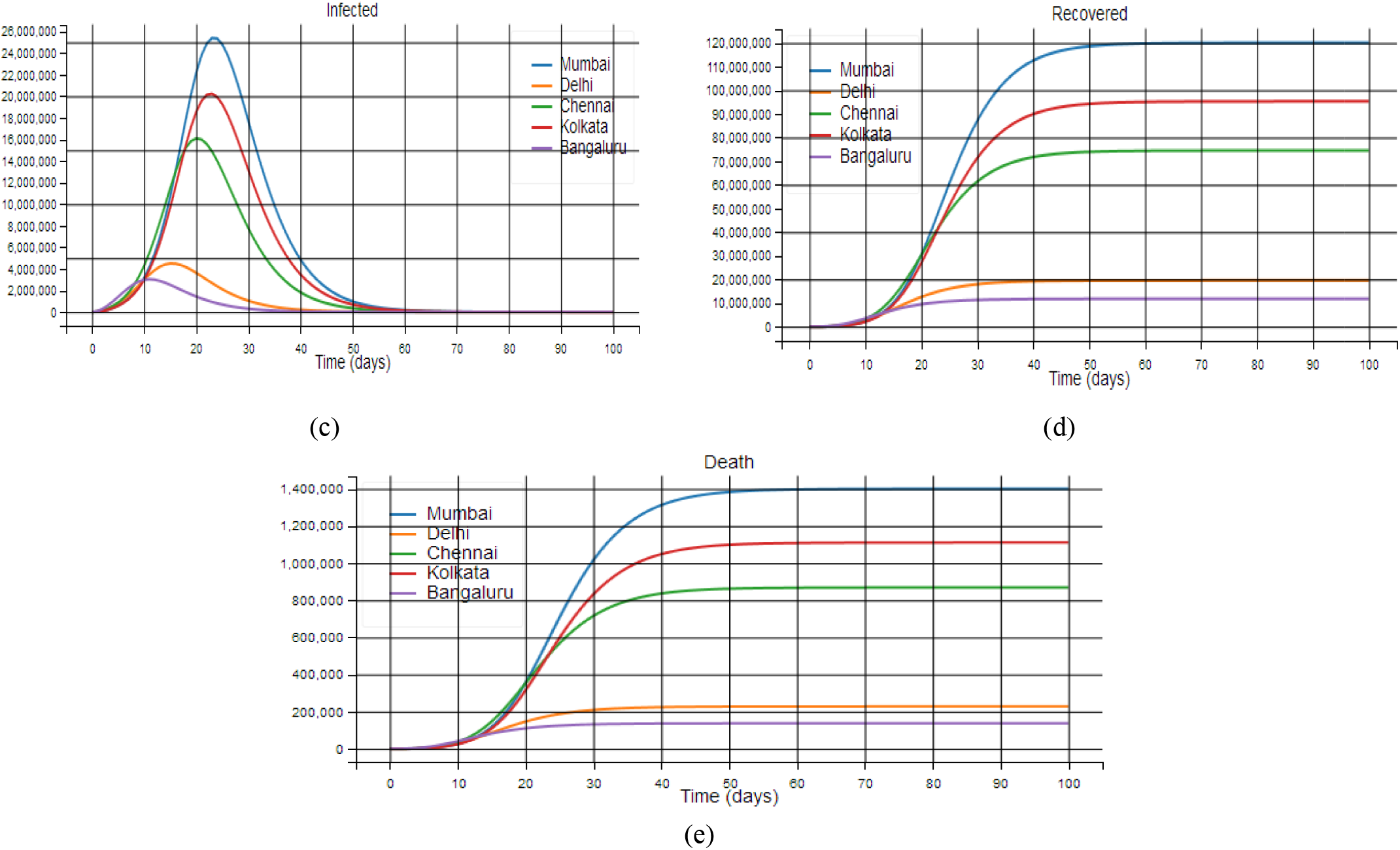
Plots showing the effect of disease transmission without lockdown in the SEIRD compartments. (a) Susceptible, (b)Exposed, (c)Infected, (d) Recovered, (e) Death

Our goal for the optimization is to reduce the infected population to the point that the economy can be kept functioning and to reduce the death toll to a minimum.

Table 2 below shows the result of the 101-day run. Since no lockdown is implemented the results are based on a full progression of the disease through the entire population with devastating consequences.

**Table 2.** Final values of S, E, I, R, and D population after 101 days the evolution of the Multicity-SEIRD Model without any lockdown implementation.

| Index | State | Population | Susceptible | Exposed | Infected | Recovered | Death | Death Rate |
| --- | --- | --- | --- | --- | --- | --- | --- | --- |
| 1 | Mumbai | 122153000 | 620291 | 8636 | 7150 | 120116150 | 1400773 | 0.011467 |
| 2 | Delhi | 19814000 | 11792 | 1607 | 1438 | 19570929 | 228234 | 0.011518 |
| 3 | Chennai | 75695000 | 263983 | 8831 | 7351 | 74545497 | 869338 | 0.011484 |
| 4 | Kolkata | 96906000 | 466193 | 7780 | 6434 | 95314057 | 1111536 | 0.011470 |
| 5 | Bengaluru | 11883000 | 16 | 12 | 15 | 11745978 | 136979 | 0.011527 |

The total death for the case of no lockdown implementation is 3746860. The decrease of bed capacity with the increase of infected population over time is given in Fig 6. It shows the dramatic lack of bed capacity in all the five states. This would be a crisis situation and was thus avoided by most countries though social distancing guidelines, lockdown and augmentation of medical resources. Hence maintaining the resource is critical component of this optimization and is accomplished through the knapsack formulation.

**Fig 6:**
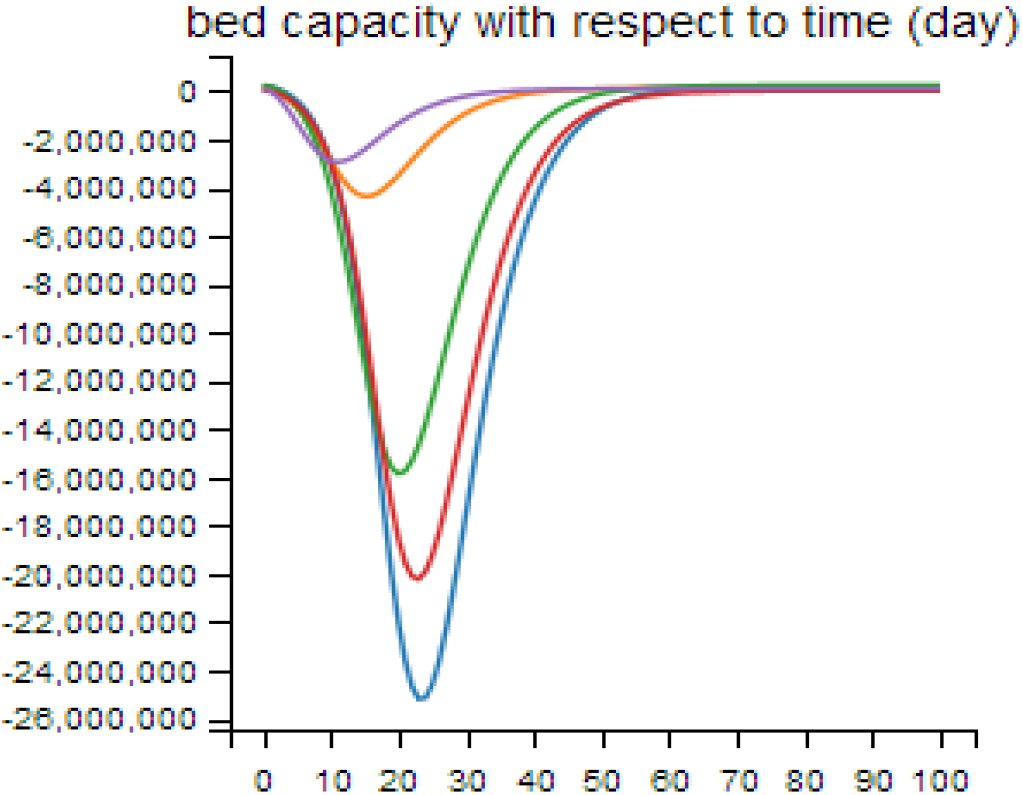
Hospital bed capacity impact without lockdown

### 3.2) Model 1: Optimal Lockdown using Quantum Knapsack Problem

We used the same initial state given in Table 1, and the same user-specified constant for the Multicity-SEIRD model as for Model 0. The time frame of 101 days is split into n = 5 days interval period, where we run the quantum knapsack problem after each interval period to figure out the optimal lockdown schedule, which is to be applied at the beginning of the next interval.

Table 3 below shows the end result of the 101-day run. Table 4 shows the lockdown schedule for each phase as determined by the optimization solution to the knapsack problem.

**Table 3.** Final values of S, E, I, R, and D population after 101 days the evolution of Multicity-SEIRD Model with optimal lockdown implementation using Quantum Knapsack Problem

| Index | State | Population | Susceptible | Exposed | Infected | Recovered | Death | Death Rate |
| --- | --- | --- | --- | --- | --- | --- | --- | --- |
| 1 | Mumbai | 122153000 | 98732786 | 3511717 | 1370505 | 18324295 | 213697 | 0.001749 |
| 2 | Delhi | 19814000 | 6986952 | 95473 | 136179 | 12450203 | 145193 | 0.007327 |
| 3 | Chennai | 75695000 | 55473016 | 2048569 | 747179 | 17225358 | 200878 | 0.002653 |
| 4 | Kolkata | 96906000 | 74775573 | 426309 | 468251 | 20991071 | 244796 | 0.002526 |
| 5 | Bengaluru | 11883000 | 568043 | 209381 | 143102 | 10836106 | 126368 | 0.010634 |

**Table 4.**
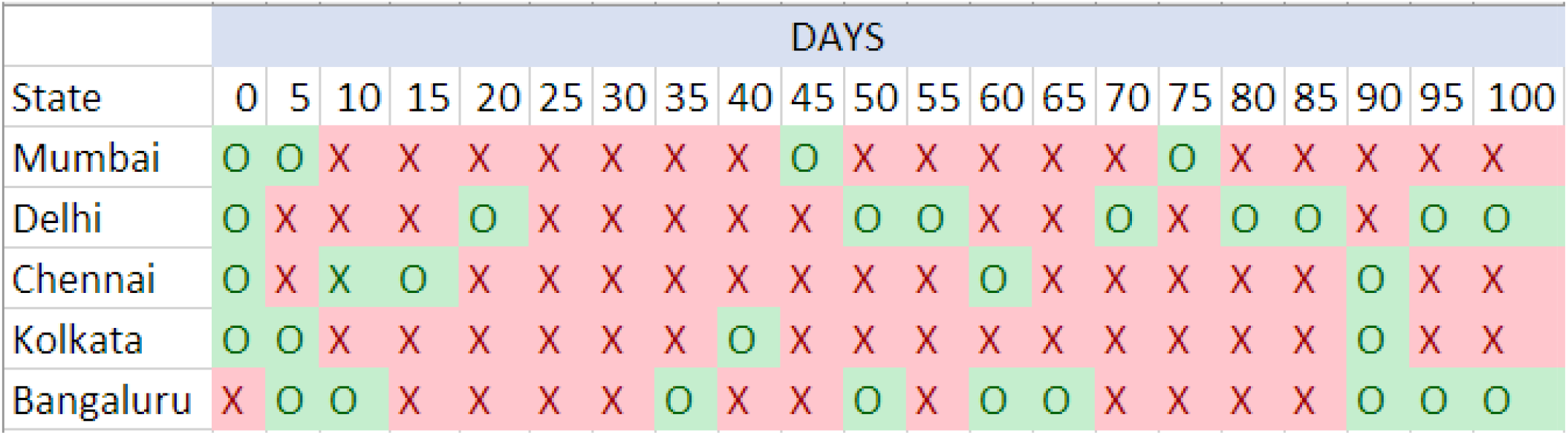
Optimal lockdown implementation using Knapsack Problem (quantum solution)

The total death by implementing an optimal lockdown schedule is 930932. The bed capacity overtime for all five states is given in Fig 7, but still there is a lack of bed availability in the hospital, so we assume the rest goes in-home quarantine. While the GDP is maintained with an average 193.25 (USD) over each Interval of 5 days.

**Fig 7:**
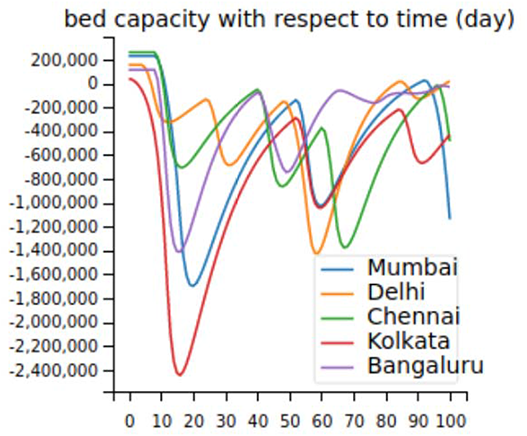
Hospital bed capacity with optimized lockdown (quantum results)

### 3.3) Model 2: Optimal Lockdown using Classical Knapsack Problem

We used the same initial state given in Table 1, and the same user-specified constant for the Multicity-SEIRD model as for Model 0. The time frame of 101 days is split into n = 5 days interval period, where we run the quantum knapsack problem after each interval period to figure out the optimal lockdown schedule, which is to be applied at the beginning of the next interval.

Table 5 shows the end result of the classical results. In addition, Table 6 shows the recommended solution of the optimal lockdown schedule for each phase.

**Table 5.** Final values of S, E, I, R, and D population after 101 days the evolution of Multicity-SEIRD Model with optimal lockdown implementation using Classical Knapsack Problem

| Index | State | Population | Susceptible | Exposed | Infected | Recovered | Death | Death Rate |
| --- | --- | --- | --- | --- | --- | --- | --- | --- |
| 1 | Mumbai | 122153000 | 90266226 | 864348 | 965623 | 29710324 | 346479 | 0.002836 |
| 2 | Delhi | 19814000 | 1848090 | 293204 | 263565 | 17208456 | 200685 | 0.010128 |
| 3 | Chennai | 75695000 | 49198915 | 1786441 | 760578 | 23672997 | 276069 | 0.003647 |
| 4 | Kolkata | 96906000 | 70739833 | 1031058 | 908102 | 23947730 | 279277 | 0.002881 |
| 5 | Bengaluru | 11883000 | 338024 | 88601 | 66222 | 11258855 | 131298 | 0.011049 |

**Table 6.**
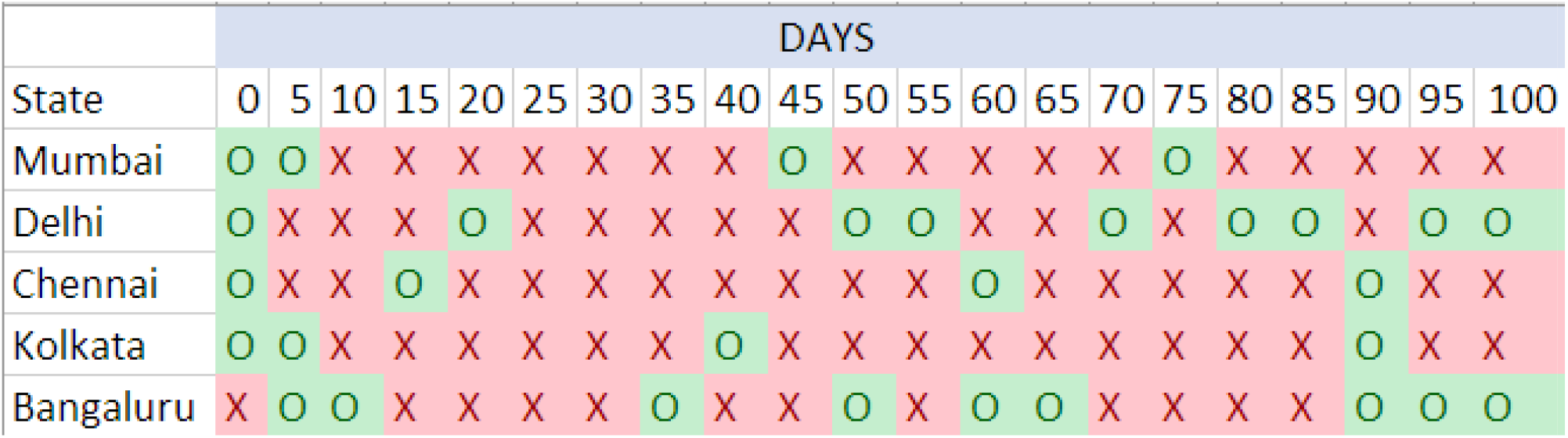
Optimal lockdown implementation using Knapsack Problem (classical solution)

The total death by implementing an optimal lockdown schedule is 1233808. The bed capacity overtime for all five states is given in Fig 8, but still there is a lack of bed availability in the hospital, so we assume the rest goes in-home quarantine. While the GDP is maintained with an average 248.24 (USD) over each Interval of 5 days.

**Fig 8:**
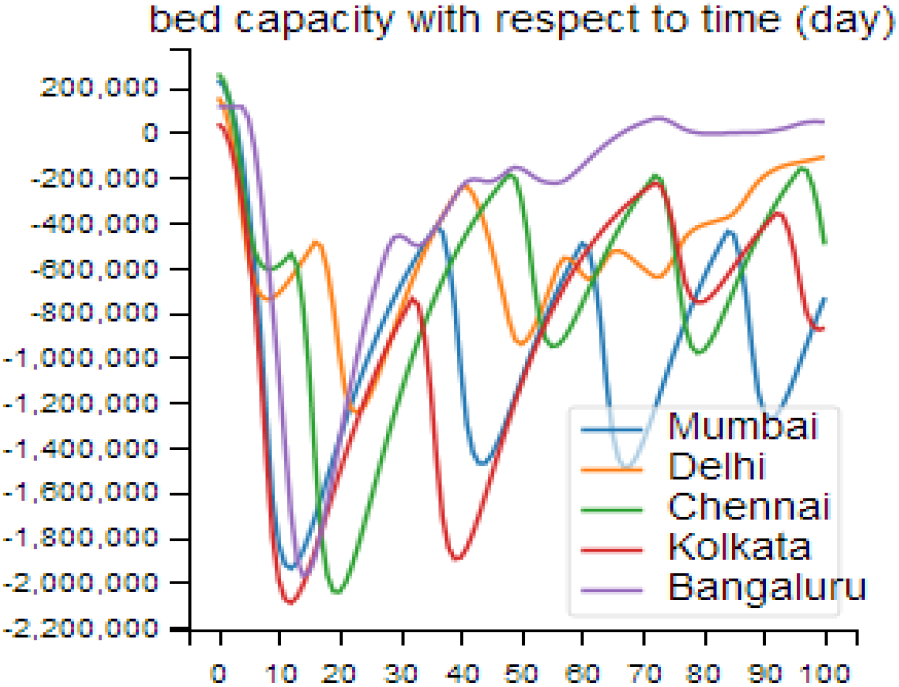
Hospital bed capacity with optimal lockdown schedule (classical results)

### 3.4) Effect of scaling

As the number of cities is increased, the QUBO requires the effect of each city to be evaluated with the other, through the transfer function. This, however, can make the problem complicated as data will most likely not be available, and validation with the actual data becomes increasingly difficult. A way around this would be to look at clusters of Cities and the smaller towns around them and make assumptions on simplifying the model. Thus, a large city with 5 surrounding cities can be modeled with Transfer functions, but only the large population center is then connected to other large cities. The effect of this simplification will have to be evaluated.

## 4. Result and Discussion

### 4.1) Case Study

We have taken a sample of 5 cities with approximate data to review the effect and effectiveness of the knapsack problem on the two objectives. An accurate representation requires more cities, evaluation of the effect of each city on each other, and calibration of the variables.

In any case, with the selected 5 cities and their sample population, we run the lockdown optimization using knapsack formulation both classically and using the quantum annealing computer D-Wave 2000Q.

**Fig 9:**
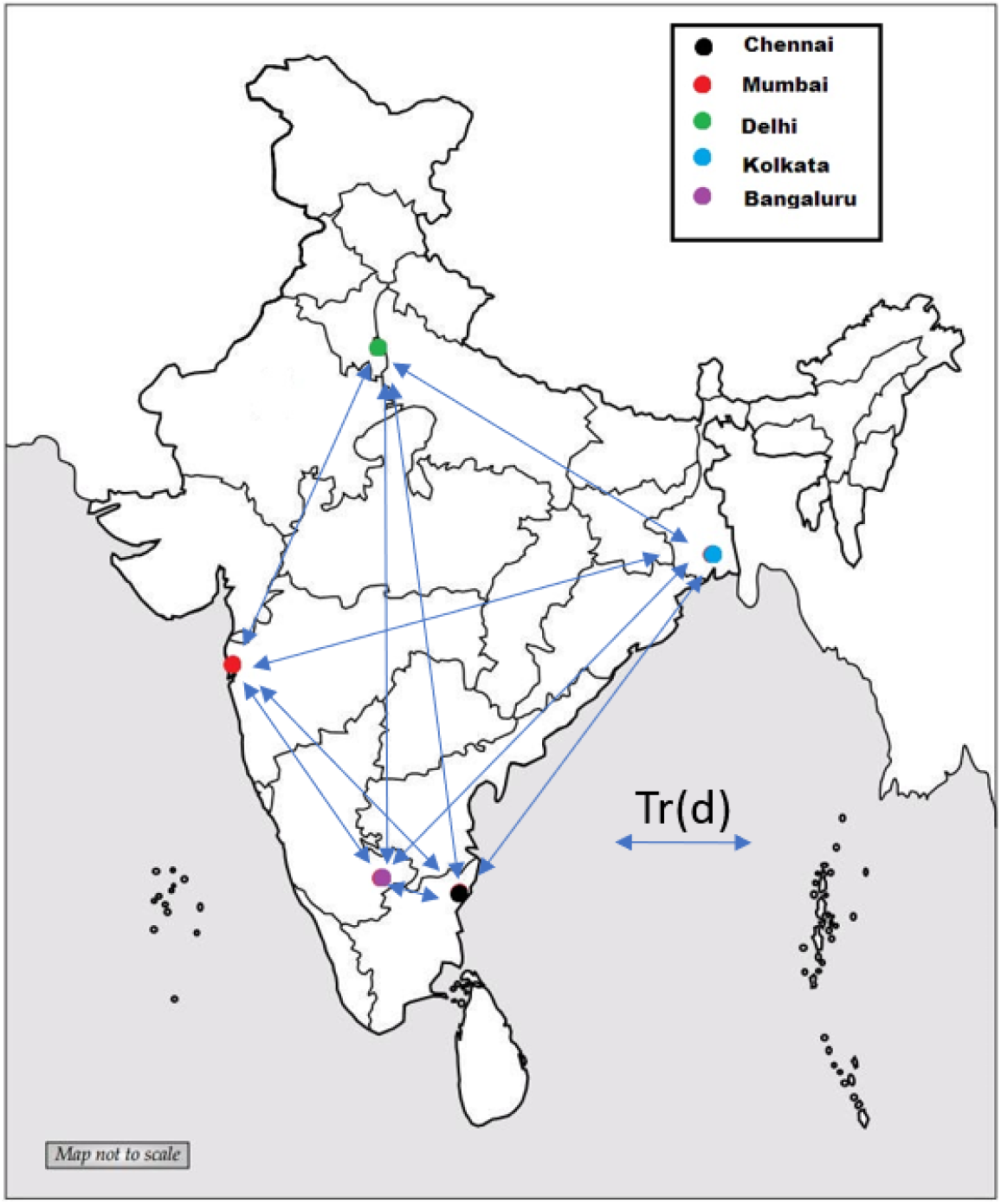
Model graph representation based on 5 regions in India with disease transmission channel along edges.

The results show that the optimization function in both cases attempted to keep cities open where the disease was not that widely spread and the infected could be accommodated in the hospitals. The remaining cities had to be shut down. The actual trend is very chaotic and impacted by many variables. Even the solutions through both the quantum and classical methods are probabilistic and thus firstly are not consistent, and secondly, small variations at each period will cause the optimization at the next period to be dramatically different.

The results in the plots below are thus only a sample and will change with each run and with actual data will look dramatically different. With that said, it can be seen that initially the GDP is pushed down, but as time goes on the Quantum solution begins to bring the GDP back up. The classical solution appears to be somewhat random causing GDP to come back up, then pushed down, and then brought all the way up before shutting back down again. This will need to be investigated further.

**Fig 10:**
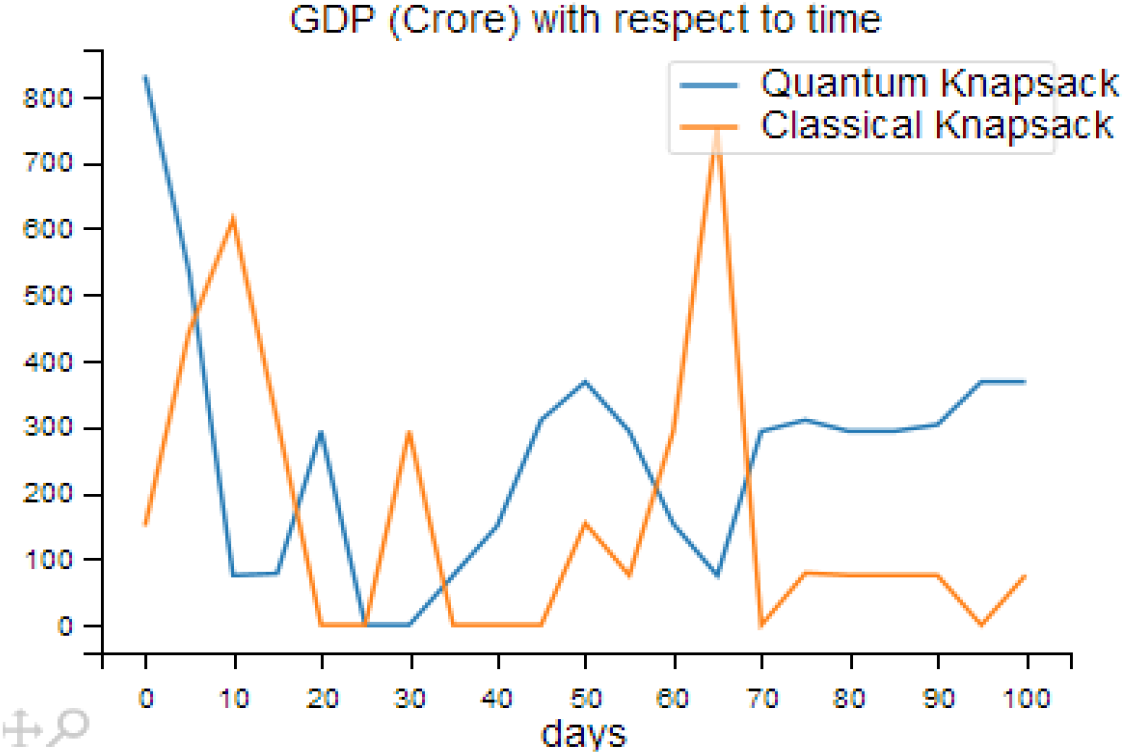
Impact of GDP through classical and quantum optimization results with time in days

In the end, it is critical that the death rate is suppressed and does not rise. It can be seen that the quantum solution also reduced the overall death rate in each city. Thus over 100 days, the plot below shows that for each city there is a dramatic reduction in the overall death count from no lockdown, but a further improvement with the solution produced from the quantum results. This has to be reviewed further since the formulation given to the quantum computer is the same as that solved classically using a probabilistic method, however, the solution from the quantum computers with very little sampling produced a superior result.

**Fig 11:**
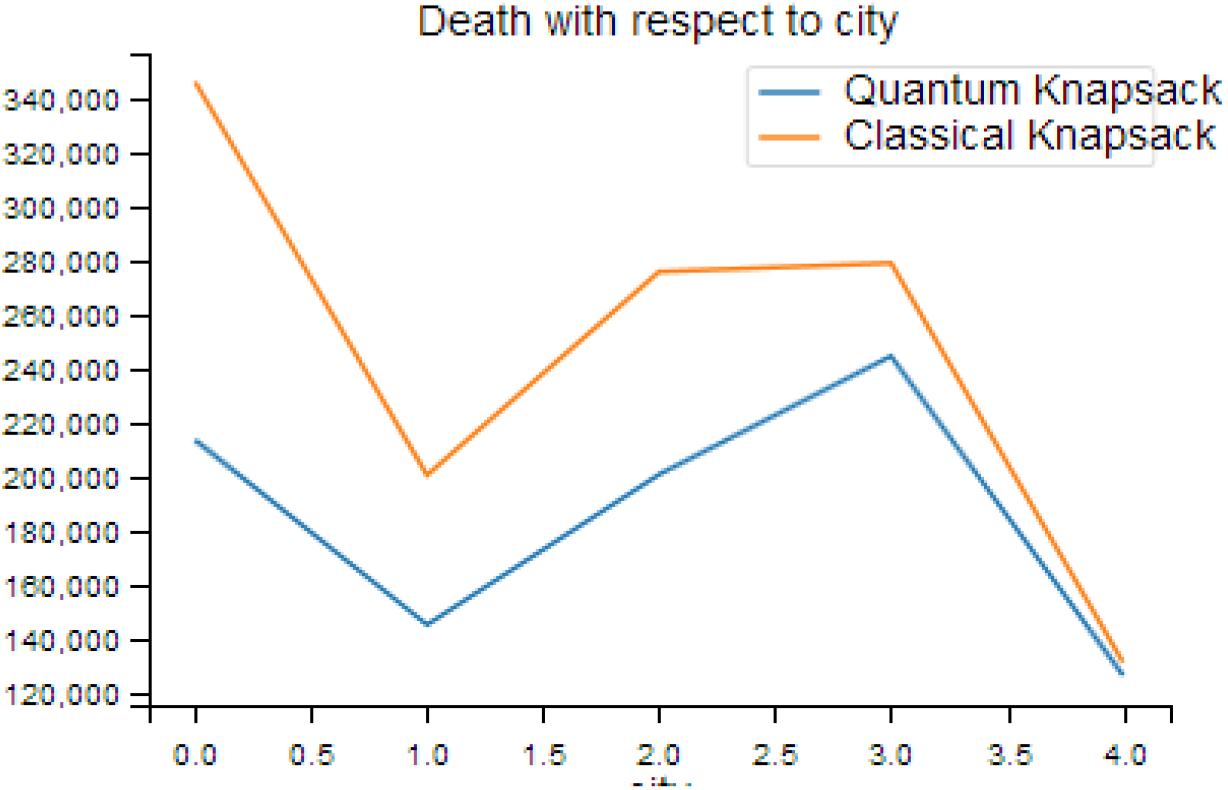
Plot showing both classical and quantum results of total death in all 5 regions (0 to 4) at the end of the 101-day period using lockdown optimization.

While managing through a highly transmissible disease, it is important to quickly identify areas of increasing infections and shut them down. This solution does more than that. It shuts down a city when the rise in infection outweighs the contribution of its GDP. It can be seen when a city is open, the infection grows, while when a city is closed, the infection begins to decline. Bengaluru is the first to be locked down due to its lower GDP contribution, however, as the infection grows in the bigger cities, they must be shut down as well. In the sample run below, Mumbai is the last to be locked down and we know it has the largest contribution to GDP. As the larger cities go into lockdown, the smaller ones are opened up, to begin to make a GDP contribution. The solutions appear to have a chaotic behavior and we assume they will change based on the run. It is also important in a real situation to monitor and adjust the parameters to fit the real-world data since the optimization must be done based on current and accurate information. The paper only brings to light that with an optimization such as this, a balancing is possible.

**Fig 12:**
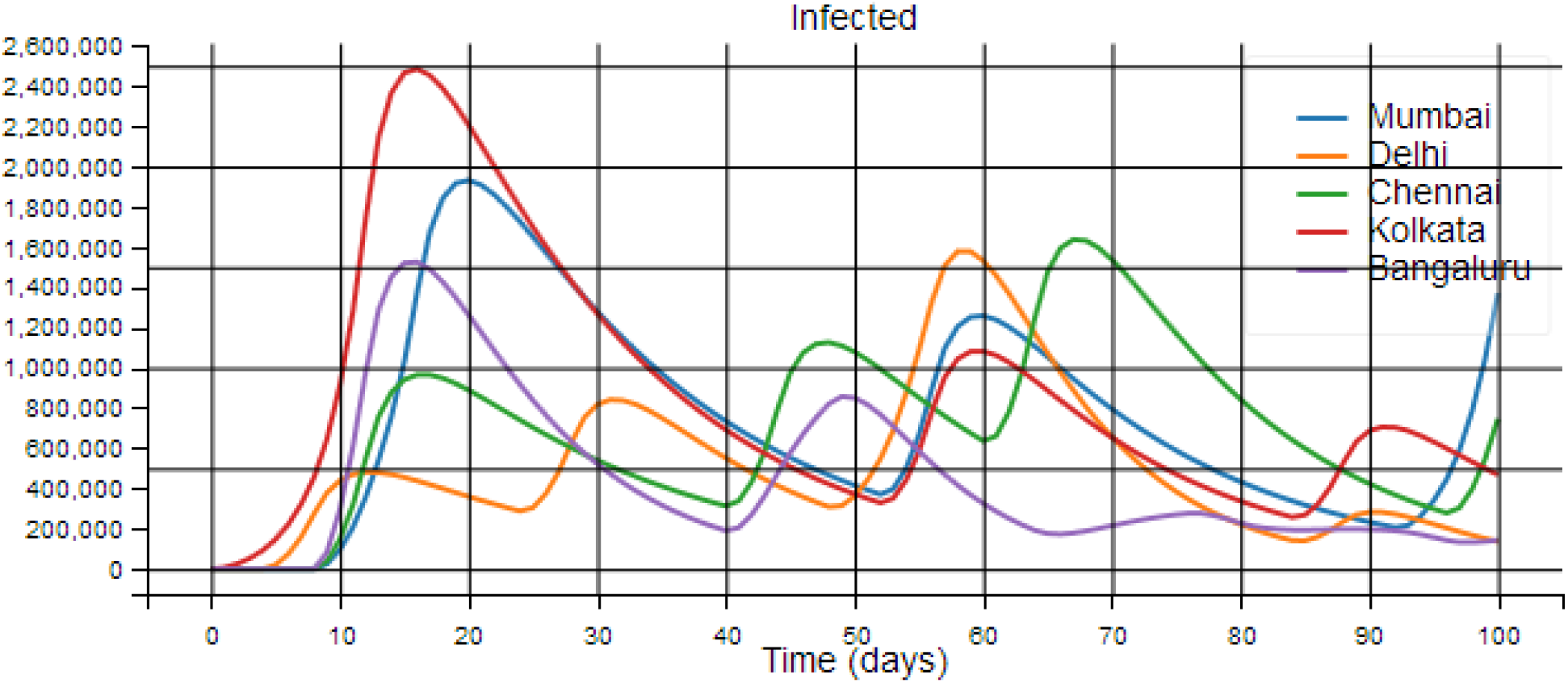
Impact of optimization on the evolution of the infected population in each region with time in days (based on the quantum results only)

### 4.2) Next Steps and Further Research

Our goal in this paper is to establish a model of optimizing the economy and addressing the impact of the disease. Therefore, we have been more concerned with whether a model can be used that accomplishes this. We have shown that a model can be created especially using a QUBO and implemented on a quantum computer which by its nature should be able to address a more complicated graph (more cities) when those are brought into the model. There are still several steps still needed to validate the model, improve on the model, and verify the model with real data. Our goals for further research include using real-world data and calibrating all the variables to match the real incident of the disease and match the actual trends in various sample cities and countries.

We see the impact and prescription of the model in reducing the impact of disease spread and also allowing the economy to open. This model needs to be compared with other methods to see if the optimization can be further improved, or whether this is truly an optimal model.

Further research based on this model includes looking at the effect of introducing vaccination. The effect of vaccination would be to reduce the susceptible population, this can be modeled to find out an optimal vaccination strategy. It can be used to see if the random introduction of vaccines is advantageous, or selective introduction based on a schedule from city to city.

One benefit of the model is to be able to predict and determine the impact of various parameters in the SEIRD model, the transmission function, or the calibration constant. Policy decisions can be made and focused on the real issues based on the impact and sensitivity of a parameter to the eventual GDP and deaths.

## 5. Conclusion

India is used as a case study in the description of the proposed updated SEIRD model integrating commerce. However, for any given country, this approach is not constrained, i.e. it can be relevant and accurate for any city or country. Our proposed SEIRD model is integrated with a transfer mechanism that helps to connect several cities or a wide region or country as a whole. This model can be extended to manage heavily populated areas like industrial campuses, schools, colleges, factories, etc. This is why the proposed model retains a clear balance between the spreading of COVID-19 and the country’s GDP (or other economic factors relevant to the organization).

While quantum computing is in its early stage, in the current and post-COVID scenarios, the optimization model we have provided can be generalized to figure out more general problems. The model will be revised in the future, when the vaccine is available, to provide a decrease in the susceptible population dependent on immunity into the model. Besides, based on the proposed model, an optimum vaccine schedule may be made. After the lockdown, our proposed model could be applied to unlock the city or town, workplace, school, colleges, universities, etc.

We show that this model can be used with the appropriate medical and policy oversight along with validation and regular calibration with real data, to meet the objective of opening up cities through the COVID-19 crisis.

## Data Availability

Data will be available on request

## Appendix

**Table 7.**
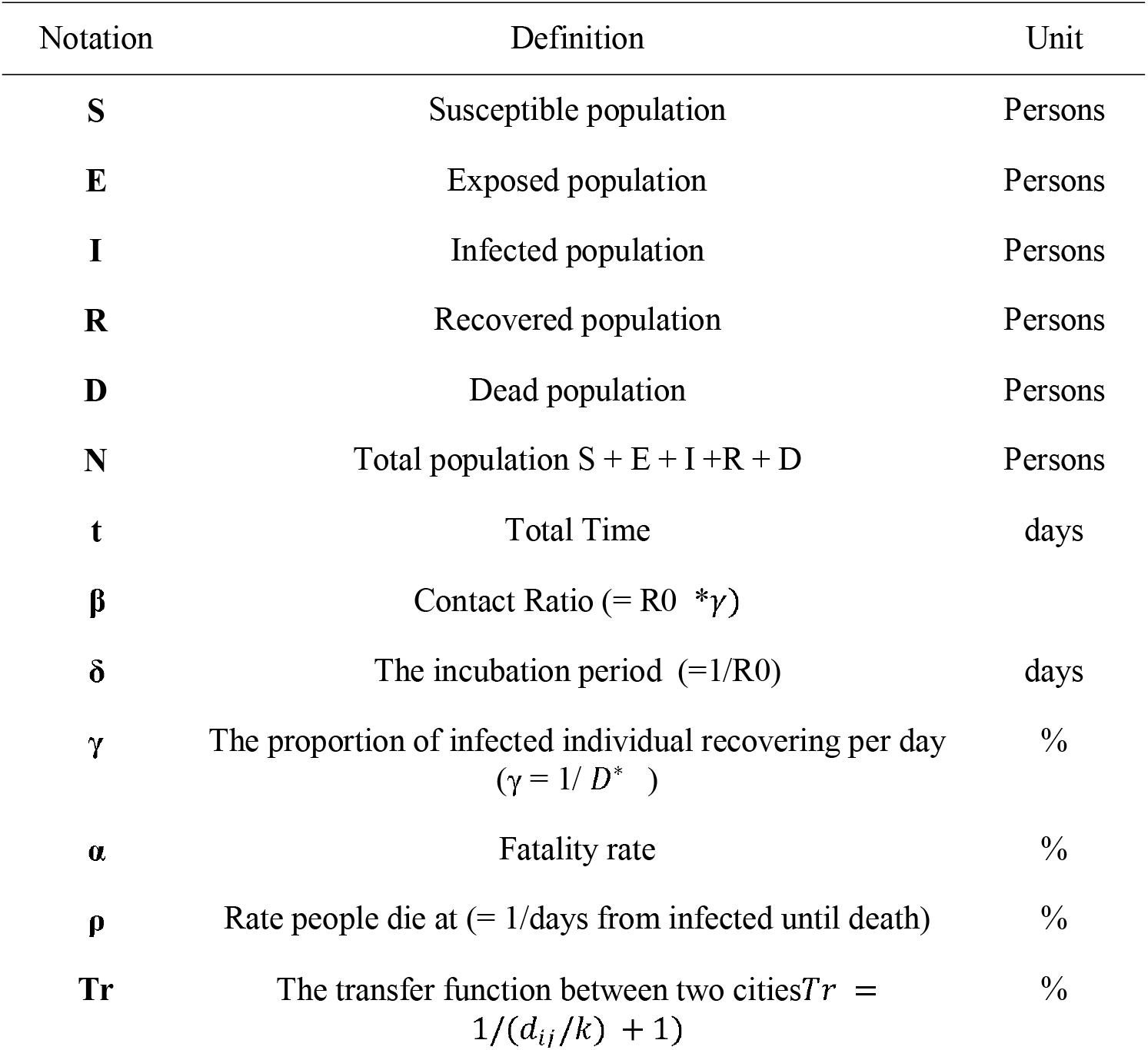
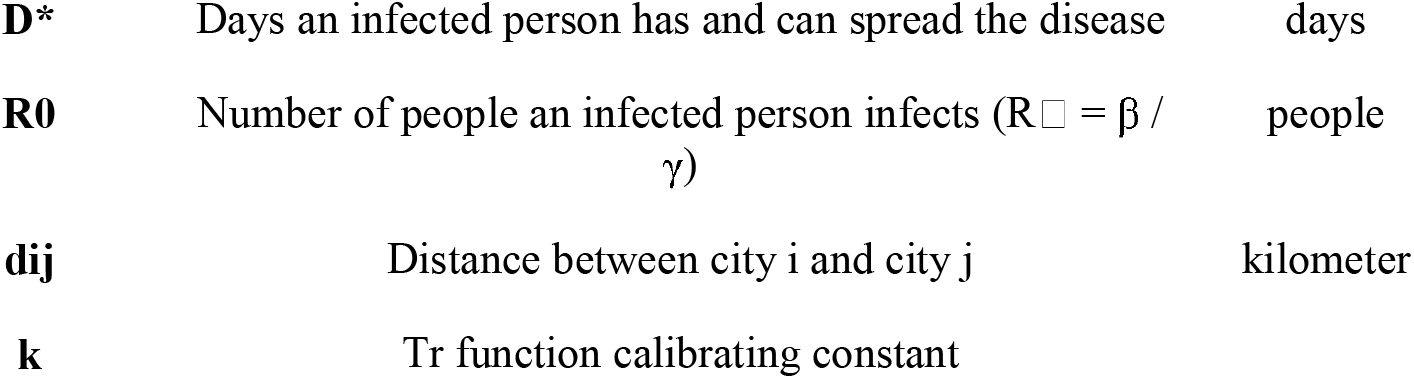
Variable and parameters, definition, unit, and values

| Notation | Definition | Unit |
| --- | --- | --- |
| **S** | Susceptible population | Persons |
| **E** | Exposed population | Persons |
| **I** | Infected population | Persons |
| **R** | Recovered population | Persons |
| **D** | Dead population | Persons |
| **N** | Total population $S + E + I + R + D$ | Persons |
| **t** | Total Time | days |
| **$\beta$** | Contact Ratio ($= R_0 * \gamma$) | |
| **$\delta$** | The incubation period ($= 1/R_0$) | days |
| **$\gamma$** | The proportion of infected individual recovering per day<br>($\gamma = 1/D^*$) | % |
| **$\alpha$** | Fatality rate | % |
| **$\rho$** | Rate people die at ($= 1/\text{days from infected until death}$) | % |
| **Tr** | The transfer function between two cities $Tr = 1/(d_{ij}/k + 1)$ | % |
| **D\*** | Days an infected person has and can spread the disease | days |
| **R0** | Number of people an infected person infects ($R_0 = \beta / \gamma$) | people |
| **dij** | Distance between city i and city j | kilometer |
| **k** | Tr function calibrating constant |  |

## 6. Acknowledgments

1. COVID formulation “Herd vs Lockdown” by Alex Khan: https://github.com/AlignedIT/SIR-Model
2. Knapsack formulation by D-Wave and Andrew Lucas: https://github.com/dwave-examples/knapsack
3. Use of Knapsack formulation for COVID by Alexander Soare in CDL-Hackathon 2020: https://github.com/CDL-Quantum/Hackathon2020/tree/master/QAlpha
4. The SEIRD model by Henry Froese: https://github.com/henrifroese/infectious_disease_modelling/blob/master/part_two.ipyn
5. We also appreciate the free use and unrestricted availability of the D-Wave 2000Q to the community for COVID research and solutions

## Notes

### Competing Interest Statement

The authors have declared no competing interest.

### Funding Statement

We appreciate the free use and unrestricted availability of the D-Wave 2000Q to the community for COVID research and solutions

### Author Declarations

IRB not required for this work

